# Prevalence of Function-Limiting Late Effects in Hodgkin Lymphoma Survivors

**DOI:** 10.1101/2020.10.02.20202408

**Authors:** Nabela Enam, Kathy Chou, Michael D. Stubblefield

## Abstract

**Objective:** To define the prevalence of neuromuscular, musculoskeletal, pain, visceral, oncologic and other late effects impacting function and quality of life in Hodgkin lymphoma(HL) survivors presenting to an outpatient cancer rehabilitation clinic.

**Design:** A retrospective cohort analysis.

**Setting:** Outpatient cancer rehabilitation clinic.

**Participants:** One hundred consecutive HL survivors.

**Interventions:** None.

**Main Outcome Measures:** The prevalence of neuromuscular, musculoskeletal, pain, visceral, oncologic and other late effects contributing to functional impairment and disability in HL survivors.

**Results:** Among the 100 HL survivors, 43% received chemotherapy, 94% radiation therapy, and 38% a combination of chemotherapy and radiation for initial treatment. Nearly all HL survivors were diagnosed with myelopathy (83%), radiculo-plexopathy (93%), mononeuropathy (95%) and localized myopathy (93%). Musculoskeletal sequelae were extremely common and included dropped head syndrome (83%), cervicalgia (79%), shoulder girdle dysfunction (73%), and dysphagia (42%). Visceral disorders were also common and included cardiovascular (70%), pulmonary (44%), endocrine (63%), gastrointestinal (29%), and genitourinary (11%) dysfunction. Lymphedema affected 21% of survivors and 30% had a history of a secondary malignancy. Pain (71%), fatigue (45%), and dyspnea (43%) were major function-limiting impairments. Nearly all (95%) of survivors were referred to at least one therapy discipline including physical therapy, occupational therapy, speech and language pathology and/or lymphedema therapy.

**Conclusion:** Neuromuscular, musculoskeletal, pain, visceral, oncologic and other late effects are extremely common in HL survivors seeking physiatric evaluation. Multiple function-limiting disorders can co-exist in HL survivors with the potential to severely compromise function and quality of life. Safe and effective rehabilitation may depend on the physiatrist’s ability to identify, evaluate and manage the multitude of complex and often interrelated functional late effects seen in HL survivors.

## Introduction

Hodgkin lymphoma (HL), formerly Hodgkin’s disease, is named for Thomas Hodgkin the physician who, in 1832, first described the clinical history and postmortem findings of massive enlargement of the lymph nodes and spleens.^1^ The first reports that x-rays could shrink the enlarged lymph nodes associated with HL emerged in the early 1900’s,^2^ and evidence that localized HL could be cured by radiation therapy (RT) was presented by Vera Peters in 1950.^3^ It was not until 1970 that HL became the first advanced malignancy of a major organ system in adults to be cured by chemotherapy.^4,5^ Combined modality therapy (CMT), meaning combining chemotherapy with RT for HL, was slow to develop, and up until the 1990’s patients were often treated with radiation as a single modality. Contemporary treatment of HL has evolved but still generally utilizes CMT with an overall 5-year survival rate of 86%.^6^ Most HL survivors treated today will enjoy an equivalent life expectancy to those of healthy age-matched individuals.

HL treatment is notorious for causing visceral, oncologic, musculoskeletal, and neuromuscular late effects.^7^ RT is particularly toxic and can damage any tissue or system within the radiation fields used to treat HL. Dysfunction is seen in the nerves and muscles encompassed by the radiation field including the spinal cord, nerve roots, plexus, peripheral nerves, and muscles. This phenomenon, termed “myelo-radiculo-plexo-neuro-myopathy,” can develop years or decades following completion of RT and progress indefinitely.^8^ Visceral late effects include those of the cardiovascular, pulmonary, gastrointestinal, urogenital, and endocrine systems. Secondary malignancies are also often seen.^9^

Considerable effort has focused on the visceral and oncologic late effects of HL survivors with studies largely occurring in oncology and survivorship settings. Resulting guidelines are generally intended for oncologists, medical survivorship specialists, and primary care physicians.

There is little data concerning the issues faced by HL survivors in the rehabilitation medicine setting.

The goal of this retrospective, cohort analysis is to identify the prevalence of neuromuscular, musculoskeletal, visceral, oncologic and other late effects contributing to functional impairment and disability in HL survivors seeking cancer rehabilitation medicine services. Comprehensively elucidating the multitude of complex and interrelated issues faced by HL survivors will educate rehabilitation physicians and improve the longitudinal care of this often misunderstood and underserved cohort of patients. Additionally, highlighting the potential impact of visceral complications on function and quality of life (QOL) in HL survivors will highlight the need for rehabilitation physicians to expand their role with respect to these disorders.

## Methods

This study was approved by the Institutional Review Board at the Kessler Foundation. Data for 100 sequential patients previously diagnosed and treated for late effects of HL was available and included in the study. Alternative primary cancer diagnoses were excluded. Each patient was seen at least once at a cancer rehabilitation outpatient clinic in New Jersey between March 2015 and March 2020. A cancer rehabilitation physiatrist evaluated the patients and chart review was completed by all authors. Data collection included patient demographics, stage of HL at diagnosis, treatment (chemotherapy and/or RT), treatment-related late effects (neuromuscular, musculoskeletal, visceral, oncologic), and rehabilitation therapy type prescribed. Treatment with chemotherapy was documented as either present or absent. Radiation fields were classified as: mantle, mediastinal, subtotal, periaortic, or total.

### Neuromuscular and Musculoskeletal Complications

Neuromuscular and musculoskeletal late effects were recorded as documented in the medical record. Clinical evaluation, including detailed history and comprehensive physical examination, was the primary basis for diagnosis of all neuromuscular and musculoskeletal disorders. When the spinal cord, nerve roots, plexus, peripheral nerves and muscles within the radiation field were affected together, it was termed “myelo-radiculo-plexo-neuro-myopathy” (this appeared on page 3).^8^ Neuromuscular complications were identified as myelopathy, radiculopathy, plexopathy, mononeuropathy or myopathy based upon the clinical criteria defined below and as previously described.^10^

- Myelopathy, usually subacute in presentation, was suspected in patients demonstrating neurologic deficits consistent with spinal cord involvement. This included loss of bowel or bladder function (including detrusor sphincter dyssynergia), sexual dysfunction, and upper motor signs (eg. hyperreflexia, clonus and spasticity).
- Radiculopathy and plexopathy, resulting from nerve root or plexus compression, respectively, were suspected in patients demonstrating sensory abnormalities in the appropriate dermatomal distribution, weakness in muscles innervated by the affected structure, and diminished deep tendon reflexes in an anatomically congruent distribution. As these conditions could not be reliably differentiated from each other based on clinical grounds when co-existent, patients exhibiting these signs and symptoms were usually grouped together as a single entity (radiculo-plexopathy) unless clinical evidence indicated one structure (i.e. plexus) was more severely affected.
- Mononeuropathy was suspected when patients presented with symptoms of numbness, tingling, burning, pain, weakness, and/or atrophy in the distribution of a peripheral nerve.
- Myopathy was suspected in the setting of primary muscle disease with or without nerve involvement and characterized by weakness with muscle wasting and atrophy often accompanied by pain. This was differentiated from atrophy due to denervation based on the clinical pattern of exaggerated muscle atrophy within the radiation field compared to muscle atrophy in the distribution of affected structures such as the nerve root, plexus, and peripheral nerves.

Other primarily neurologic complications included sexual dysfunction, cognitive impairment (e.g. memory deficits) and gait dysfunction.

Although primary bone, joint, and soft tissue dysfunction underlie the musculoskeletal disorders encountered in HL survivors, neuromuscular damage is likely the major contributing factor. While some disorders, such as dropped head syndrome (DHS) and dysphagia are usually considered neuromuscular in etiology, the effects of radiation on soft tissue and bone (vertebrae, hyoid) are likely important in HL survivors and support their classification as at least partially musculoskeletal for the purposes of this study. Musculoskeletal syndromes resulting from neuromuscular damage in HL survivors include DHS, cervicalgia, shoulder girdle dysfunction (SGD), dysphagia, and trismus with clinical criteria for diagnosis defined below.

- DHS was defined by weakness in head elevation that ranged from mild (end-of-day cervical fatigue and pain) to severe (minimal or no neck extension, muscle bulk or strength). This diagnosis was based on self-reported history and/or physical examination wherein the patient demonstrated difficulty elevating the head with absent or feeble cervical paraspinal muscle contraction on palpation.
- Cervicalgia was defined in patients reporting neck pain with a history of radiation exposure with or without evidence of DHS.
- SGD was defined as limited shoulder range of motion, weakness in active movement, and/or pain. Rotator cuff tendonitis, adhesive capsulitis, osteoarthritis, and other specific shoulder pathology diagnoses were consolidated into SGD.
- Dysphagia was defined by patient complaints of difficulty swallowing, choking episodes, or diagnosed from formal swallow evaluation.
- Trismus was defined as contraction of the muscles and other tissues of mastication leading to restricted jaw opening.

### Visceral and Oncologic Complications

Visceral and oncologic complications seen in HL survivors included cardiac, pulmonary, endocrine, gastrointestinal, genitourinary dysfunction, lymphedema, and secondary malignancies. These complications were recorded based upon outside physician consultation and documentation, patient’s self-reporting, or suspicion and investigation by the physiatrist.

- Cardiovascular dysfunction included documented history of valvular dysfunction, arrhythmias, coronary artery disease, pericardial dysfunction, congestive heart failure, myocardial dysfunction, carotid stenosis and baroreceptor failure. Baroreceptor failure was characterized by fluctuations in blood pressure and heart rate resulting from damage to the receptors (carotid baroreceptors) in the neck.
- Pulmonary dysfunction included documented pulmonary fibrosis based on restrictive lung disease pattern confirmed on pulmonary function tests.
- Endocrine dysfunction included documented history of hypothyroidism or osteoporosis/osteopenia.
- Gastrointestinal dysfunction included documented history of dyspepsia (i.e. ulcers and reflux) or bowel dysmotility.
- Genitourinary dysfunction included documented history of detrusor sphincter dyssynergia, urinary retention, or incontinence.
- Lymphedema was included when documented in the patient’s history or observed on physiatric evaluation.
- Secondary malignancy included documented history of any cancer following initial diagnosis of HL deemed likely secondary to HL treatment.

### Functional Impairments

The presence of symptoms that interfered with QOL and independence in activities of daily living (ADLs) were documented. This included patient reports of pain, fatigue, and dyspnea.

### Therapy Referrals

The type of therapy prescribed including physical therapy (PT), occupational therapy (OT), speech language pathology therapy (SLP), and lymphedema therapy was recorded. Patients receiving multiple therapy disciplines were noted.

## Results

Charts of 100 HL survivors were reviewed (Table 1). Of these, 63 (63%) were female. Twenty-two (22%) were diagnosed with HL before age 19, 51% were diagnosed between 20-29 years of age, 19% were diagnosed between 30-39 years of age, and 8% were diagnosed over the age of 40. Known cancer staging at the time of initial HL diagnosis showed 17 (17%) patients with stage I, 40 (40%) with stage II, 19 (19%) with stage III, and 5 (5%) with stage IV (Table 1). Twenty-four survivors had nodular sclerosing HL, 1 had mixed cellularity type, 1 had both, and 74 had an unknown cellular type (data not shown).

**Table 1.** Hodgkin Lymphoma survivor demographics, lymphoma history and treatment

| **Variable** | **N (%)** |
| --- | --- |
| Sex |  |
| Men | 37 (37%) |
| Women | 63 (63%) |
| Age at Diagnosis |  |
| < 19yrs | 22 (22%) |
| 20-29yrs | 51 (51%) |
| 30-39yrs | 19 (19%) |
| 40-49yrs | 5 (5%) |
| > 50yrs | 3 (3%) |
| Hodgkin's Stage at Diagnosis |  |
| Stage 1A, 1B, I unspecified | 12 (12%), 3 (3%), 2 (2%) |
| Stage IIA, IIB, II unspecified | 24 (24%), 12 (12%), 4 (4%) |
| Stage IIIA, IIIB, III unspecified | 10 (10%), 5 (5%), 4 (4%) |
| Stage IVA, IVB, IV unspecified | 1 (1%), 2 (2%), 2 (2%) |
| Unknown stage | 19 (19%) |
| Received chemotherapy | 43 (43%) |
| No chemotherapy | 55 (55%) |
| Unknown chemotherapy history | 2 (2%) |
| Radiation treatment | 94 (94%) |
| Mantle | 44 (44%) |
| Mediastinal | 2 (2%) |
| Subtotal | 6 (6%) |
| Periaortic | 2 (2%) |
| Total | 35 (35%) |
| Unknown | 5 (5%) |
| No radiation treatment | 6 (6%) |

Among all survivors, 43 (43%) received chemotherapy and 94 (94%) received radiation (Table 1). Thirty-eight (38%) received combined chemotherapy and radiation as their initial HL treatment (data not shown). The majority of survivors received radiation involving the thoracic region, 44 (44%) received mantle, 2 (2%) isolated mediastinal, 6 (6%) subtotal, 2 (2%) periaortic, and 35 (35%) total nodal radiation (Table 1). The known radiation total doses ranged from 1400 cGy to 4920 cGy.

The majority of HL survivors demonstrated signs and symptoms consistent with myelo-radiculo-plexo-neuro-myopathy. Eighty-three (83%) demonstrated myelopathy, 93% radiculo-plexopathy, 95% mononeuropathy, and 93% myopathy. Of the neurologic complications, mononeuropathy was most common, manifesting in 95 (95%) of survivors. All subjects demonstrated at least one concurrent neurologic complication. Other neurologic complications identified in HL survivors included 8% with sexual dysfunction, 11% with neurocognitive impairments, and 39% with gait dysfunction (Table 2).

**Table 2.** Diagnoses identified in Hodgkin Lymphoma survivors

| **Diagnosis** | **N (%)** |
| --- | --- |
| Neuromuscular |  |
| Myelopathy | 83 (83%) |
| Radiculo-plexopathy | 93 (93%) |
| Mononeuropathy | 95 (95%) |
| Myopathy | 93 (93%) |
| Other Neurologic |  |
| Sexual Dysfunction | 8 (8%) |
| Cognitive Impairment | 11 (11%) |
| Gait Dysfunction | 39 (39%) |
| Musculoskeletal |  |
| Dropped head syndrome | 83 (83%) |
| Cervicalgia | 79 (79%) |
| Shoulder girdle dysfunction | 73 (73%) |
| Dysphagia | 42 (42%) |
| Trismus | 3 (3%) |
| Visceral |  |
| Cardiovascular dysfunction | 70 (70%) |
| 2 cardiovascular impairments | 15 (15%) |
| 3 cardiovascular impairments | 7 (7%) |
| 4 cardiovascular impairments | 2 (2%) |
| Valvular | 43 (43%) |
| Arrhythmia | 27 (27%) |
| Coronary artery disease | 13 (13%) |
| Pericardial dysfunction | 7 (7%) |
| Congestive heart failure | 6 (6%) |
| Myocardial dysfunction | 4 (4%) |
| Baroreceptor Failure | 4 (4%) |
| Carotid stenosis | 3 (3%) |
| Pulmonary fibrosis | 44 (44%) |
| Endocrine dysfunction | 63 (63%) |
| Hypothyroidism | 59 (59%) |
| Osteopenia/osteoporosis | 17 (17%) |
| Gastrointestinal dysfunction | 29 (29%) |
| Dyspepsia | 17 (17%) |
| Bowel dysmotility | 12 (12%) |
| Genitourinary dysfunction | 11 (11%) |
| Lymphedema | 21 (21%) |
| Oncologic | 52 (52%) |
| Secondary malignancy | 30 (30%) |
| Cancer Recurrence | 22 (22%) |
| Functional impairment |  |
| Pain | 71 (71%) |
| Fatigue | 45 (45%) |
| Dyspnea | 43 (43%) |

Musculoskeletal clinical syndromes resulting from neuromuscular dysfunction were common in HL survivors including 83 (83%) with DHS, 73 (73%) with SGD, 79 (79%) with cervicalgia, 42 (42%) with dysphagia and 3 (3%) with trismus (Table 2).

The cardiovascular system was the second most commonly affected system with 70 (70%) experiencing at least one cardiovascular disorder and 15 (15%) two, 7 (7%) three and 2 (2%) four cardiovascular impairments. Of these, 43 (43%) had valvular abnormalities, 27 (27%) cardiac arrhythmias, 13 (13%) coronary artery disease, 7 (7%) pericardial dysfunction, 6 (6%) congestive heart failure, 4 (4%) myocardial dysfunction, 4 (4%) baroreceptor failure, and 3 (3%) carotid stenosis (Table 2).

Pulmonary fibrosis was diagnosed in 44 (44%) patients, and 21 (21%) experienced lymphedema (Table 2). Fifty-nine (59%) survivors were diagnosed with hypothyroidism and 17 (17%) with osteoporosis or osteopenia. A total of 29 (29%) patients experienced gastrointestinal dysfunction including 12 (12%) with dysmotility and 17 (17%) with dyspepsia. Eleven (11%) survivors reported genitourinary abnormalities (Table 2).

HL recurrence occurred in 22 (22%) patients and 30 (30%) patients developed secondary malignancies. Forty-three (43%) patients described dyspnea, 45 (45%) experienced fatigue, and 71 (71%) reported pain (Table 2).

Therapy was prescribed to the vast majority (95%) of survivors (Table 3). Twenty-eight (28%) required two disciplines, 7 (7%) three disciplines, and 1 (1%) four disciplines. Ninety-four (94%) survivors received PT, 15 (15%) received OT, 21 (21%) SLP, and 12 (12%) received lymphedema therapy.

**Table 3.** Therapy referrals for Hodgkin Lymphoma survivors

| **Therapy Referrals** | **N (%)** |
| --- | --- |
| Any | 95 (95%) |
| 2 disciplines | 28 (28%) |
| 3 disciplines | 7 (7%) |
| 4 disciplines | 1 (1%) |
| Physical therapy | 94 (94%) |
| Speech language pathology | 21 (21%) |
| Occupational therapy | 15 (15%) |
| Lymphedema therapy | 12 (12%) |

## Discussion

HL is a highly curable disease owing to advances in treatment. The cost of cure, however, is extremely high for the survivors who develop severe function-limiting late effects (Figure 1a, 1b, 1c). These impairments encompass neuromuscular, musculoskeletal, pain, visceral, and oncologic abnormalities. Unchallenged by intervention, function and QOL will decline indefinitely, a fact that has led radiation late effects to be labeled with the moniker “the gift that keeps on giving”. Although more work clarifying the value of specific rehabilitation interventions is needed, the fundamental role for physiatry in identifying and managing the functional late effects common to HL survivors is clear.

**Figure 1.**
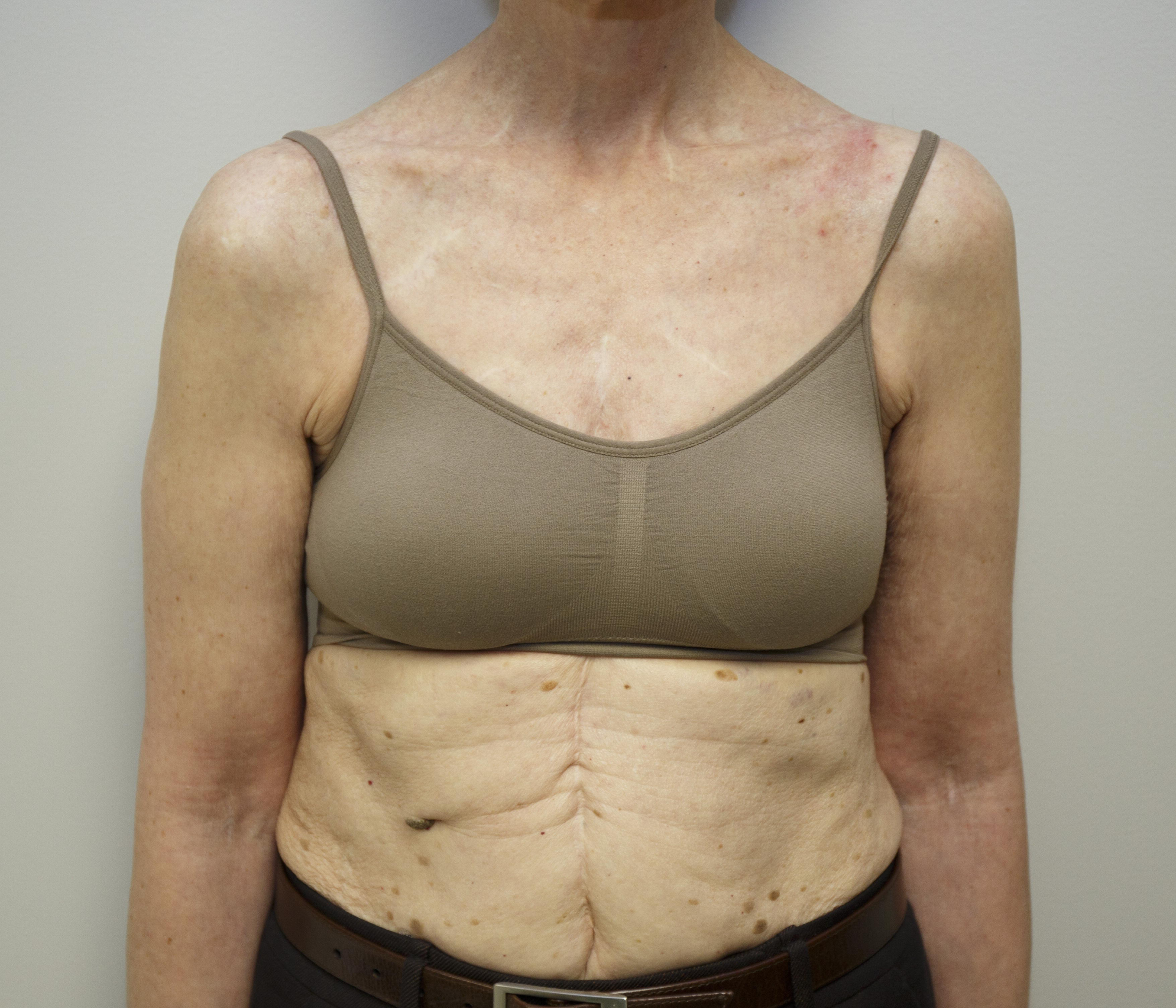
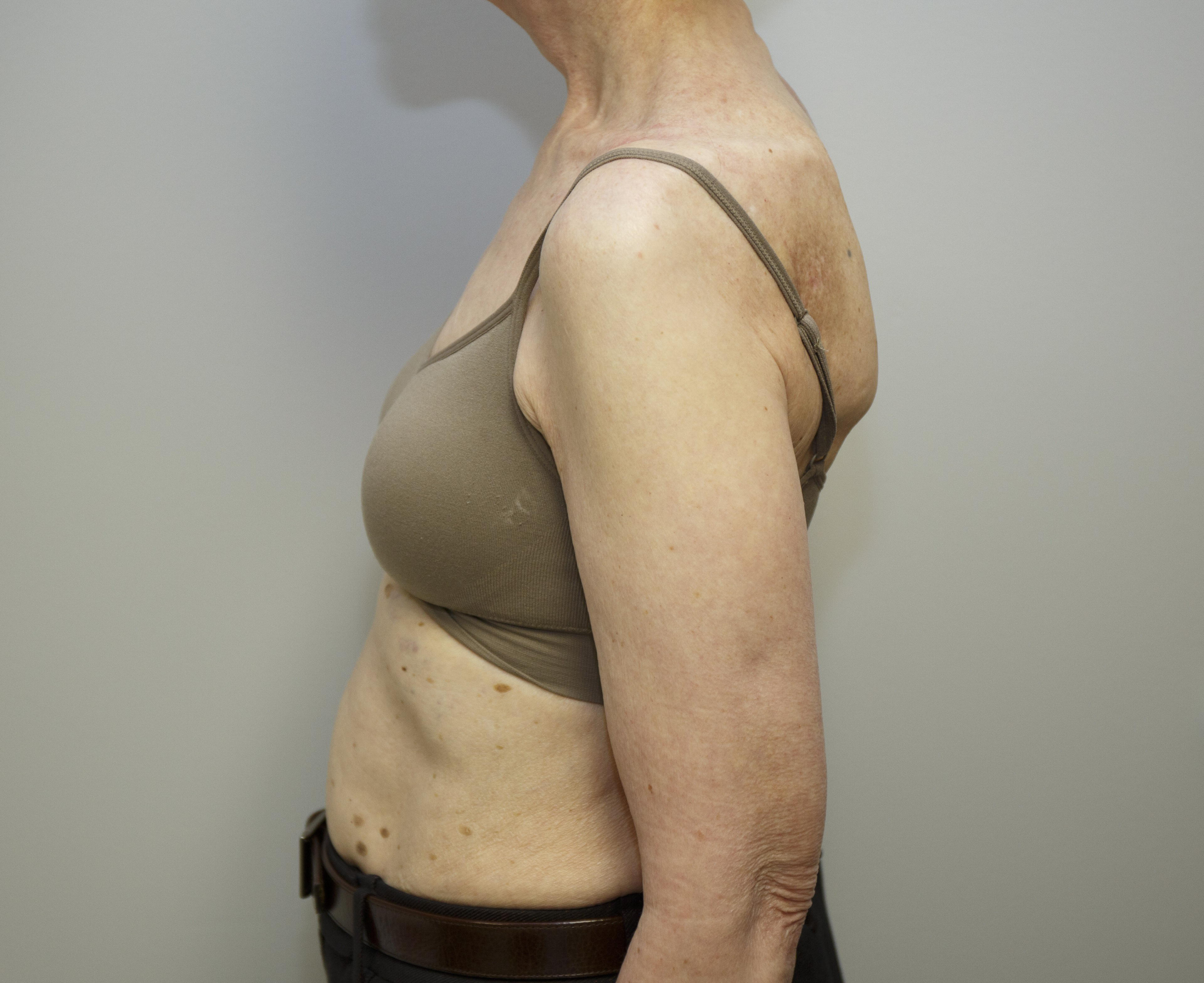
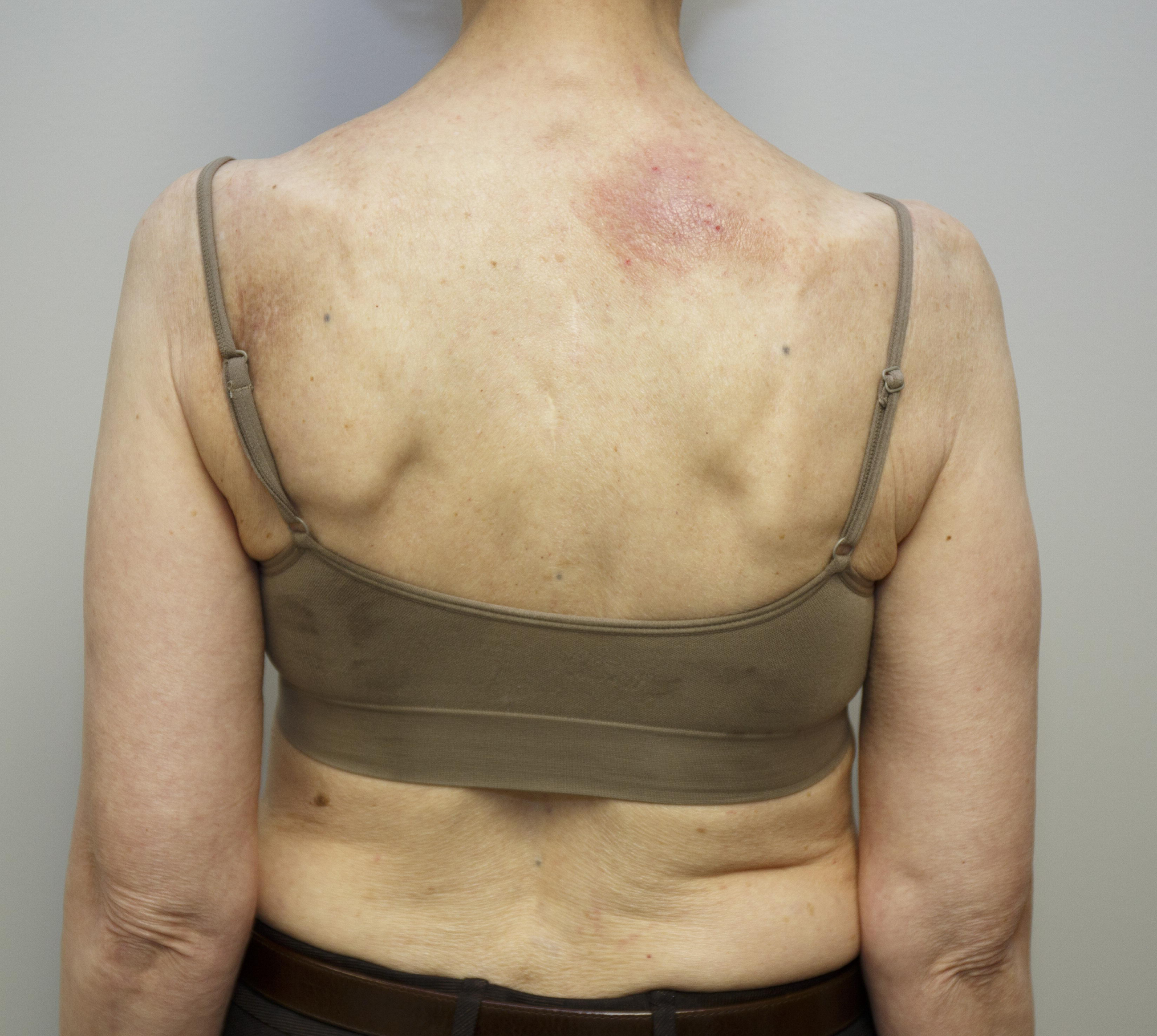
A typical Hodgkin lymphoma survivor. This woman in her 60’s has a history of stage II nodular sclerosing Hodgkin lymphoma that was diagnosed in her 30’s. She was treated with subtotal radiation resulting in multiple neuromuscular (myelopathy, radiculopathy, plexopathy, mononeuropathy, myopathy), musculoskeletal (dropped head syndrome, cervicalgia, shoulder dysfunction), functional (pain, fatigue, dyspnea) and visceral (cardiovascular, pulmonary, endocrine) late effects.

In our chart review of 100 HL survivors referred to a cancer rehabilitation specialist, all had one or more function limiting issues as would be expected based on their seeking physiatric evaluation and treatment. Most patients had multiple impairments. This is consistent with other studies that have identified the systemic late effects likely to be seen in this population.^11,12^ Prior investigations have not, however, focused on the neuromuscular, musculoskeletal, and functional complications that are likely to affect QOL and ADL in HL survivors.

Neuromuscular deficits can affect all levels of the neural axis in HL survivors. Nearly all (95%) survivors exhibited at least one neuromuscular disorder and most (83%) were found to have multiple levels affected based on clinical evaluation. Neuromuscular impairments in this population are usually attributed to radiation-induced damage of nerves and muscles within or traversing the radiation field, a phenomenon termed “myelo-radiculo-plexo-neuro-myopathy”.^8,10^ Disorders such as radiculopathy from degenerative causes and peripheral neuropathy from diabetes or neurotoxic chemotherapy exposure are also potential factors in some patients. The musculoskeletal syndromes observed included DHS (83%), cervicalgia (79%), SGD (73%), and dysphagia (42%) which is consistent with radiation-induced impairments observed in other cohorts.^13^ It is the authors opinion that the musculoskeletal syndromes described in this study result in large part from, and are secondary to, neuromuscular dysfunction which should be considered the primary disorder.

Cardiovascular complications were the second most prevalent following neuromuscular disorders, affecting 70% of survivors. Previous studies have demonstrated that radiation in HL survivors is associated with autonomic dysfunction^14^, valvular dysfunction^15^, and that cardiac mortality is a leading cause of death.^16^ A significant number of survivors had pulmonary (44%) and endocrine (63%) disorders as well. Twenty-two percent had a previously documented recurrence of HL and 30% developed a secondary malignancy. The high prevalence of visceral impairment in this study should serve to emphasize the tremendous complexity and need for close medical management of this population.

The importance of screening in HL survivors is increasingly recognized. Surveillance guidelines for late effects have been published. Particular attention has been given to cardiovascular and secondary malignancies which confer significant mortality in this population.^11,17^ The National Comprehensive Cancer Network (NCCN) as well as many societies have established recommended follow-up and surveillance algorithms for HL survivors. These guidelines include regular bloodwork, thyroid evaluation, cardiovascular risk and symptom assessment, and regular cancer screening.^9,18,19^

There are multiple treatments available that may improve function and optimize QOL in HL survivors. For physiatrists, this includes controlling pain, prescribing appropriate therapeutic interventions (i.e., PT, OT, SLP), and monitoring for changes in clinical status. Because of the tremendous medical complexity associated with HL survivors, coordination of care with other medical disciplines such as oncology, cardiology, and endocrinology is critically important for optimal rehabilitation outcomes. Physiatrists should seek to understand the full scope and scale of medical issues affecting HL survivors as they are instrumental to identifying issues early and coordinating care appropriately.

### Study Limitations

There are several limitations to our study. The high percentage of patients demonstrating signs and symptoms of myelo-radiculo-plexo-neuro-myopathy and resulting neuromuscular complications likely represents a highly biased population of patients seeking specialized physiatric care. This is probably not representative of all HL survivors. The neurologic diagnoses were generally based on clinical evaluation alone and may be erroneous in some cases. No attempt was made to correlate treatment with impairments or impairments with functional outcome. Future studies should seek to make these correlations and shed light on which interventions are most likely to be effective at restoring function and QOL to HL survivors.

## Conclusion

HL survivors are well known to develop neuromuscular, musculoskeletal, pain, visceral, and oncologic late effects as a result of treatment. These impairments can have major implications for function and QOL. These disorders are not only extremely common, but are likely to occur together in patients seeking cancer rehabilitation services. Physiatric evaluation and management can be instrumental in maximizing function and QOL in this cohort of cancer survivors. To achieve this potential, it is imperative that physiatrists treating these patients understand the full scope and scale of these disorders to ensure they support safe and effective rehabilitation.

## Data Availability

Data for 100 sequential patients previously diagnosed and treated for late effects of Hodgkin Lymphoma was available and included in the study

## Presentation Acknowledgements

None

## Financial Acknowledgements

None

## List of Abbreviations

ADL: activities of daily living
CMT: combined modality therapy
DHS: dropped head syndrome
HL: Hodgkin lymphoma
OT: occupational therapy
PT: physical therapy
RT: radiation therapy
SGD: shoulder girdle dysfunction
SLP: speech and language pathology therapy
QOL: quality of life

## References

1. Hodgkin. On some Morbid Appearances of the Absorbent Glands and Spleen. Med Chir Trans. 1832;17:68–114.

2. Pusey W. Cases of sarcoma and of Hodgkin’s disease treated by exposures to X-rays - a preliminary report. JAMA. 1902;38(3):166–169.

3. Peters MV. A study of survivals in Hodgkin’s disease treated radiologically. Am J Roentgenol. 1950;63(3):299–311.

4. Devita VT, Jr., Serpick AA, Carbone PP. Combination chemotherapy in the treatment of advanced Hodgkin’s disease. Ann Intern Med. 1970;73(6):881–895.

5. Canellos GP, Rosenberg SA, Friedberg JW, Lister TA, Devita VT. Treatment of Hodgkin lymphoma: a 50-year perspective. J Clin Oncol. 2014;32(3):163–168.

6. Cancer Stat Facts: Hodgkin Lymphoma. National Cancer Institute. https://seer.cancer.gov/statfacts/html/hodg.html.

7. Ng AK, van Leeuwen FE. Hodgkin lymphoma: Late effects of treatment and guidelines for surveillance. Seminars in hematology. 2016;53(3):209–215.

8. Stubblefield MD. Radiation fibrosis syndrome: neuromuscular and musculoskeletal complications in cancer survivors. PM R. 2011;3(11):1041–1054.

9. Darrington DL, Vose JM. Appropriate surveillance for late complications in patients in remission from Hodgkin lymphoma. Curr Hematol Malig Rep. 2012;7(3):200–207.

10. Stubblefield MD. Neuromuscular complications of radiation therapy. Muscle Nerve. 2017;56(6):1031–1040.

11. Ng AK. Current survivorship recommendations for patients with Hodgkin lymphoma: focus on late effects. Blood. 2014;124(23):3373–3379.

12. Kilickap S, Barista I, Ulger S, et al. Long-term complications in Hodgkin’s lymphoma survivors. Tumori. 2012;98(5):601–606.

13. Ghosh PS, Milone M. Clinical and laboratory findings of 21 patients with radiation-induced myopathy. J Neurol Neurosurg Psychiatry. 2015;86(2):152–158.

14. Groarke JD, Tanguturi VK, Hainer J, et al. Abnormal exercise response in long-term survivors of hodgkin lymphoma treated with thoracic irradiation: evidence of cardiac autonomic dysfunction and impact on outcomes. J Am Coll Cardiol. 2015;65(6):573–583.

15. Bijl JM, Roos MM, van Leeuwen-Segarceanu EM, et al. Assessment of Valvular Disorders in Survivors of Hodgkin’s Lymphoma Treated by Mediastinal Radiotherapy +/- Chemotherapy. Am J Cardiol. 2016;117(4):691–696.

16. Jaworski C, Mariani JA, Wheeler G, Kaye DM. Cardiac complications of thoracic irradiation. J Am Coll Cardiol. 2013;61(23):2319–2328.

17. Thompson CA, Mauck K, Havyer R, Bhagra A, Kalsi H, Hayes SN. Care of the adult Hodgkin lymphoma survivor. Am J Med. 2011;124(12):1106–1112.

18. Engert A, Dreyling M, Group EGW. Hodgkin’s lymphoma: ESMO clinical recommendations for diagnosis, treatment and follow-up. Ann Oncol. 2008;19 Suppl 2:ii65–66.

19. MD Anderson Cancer Center. Survivorship-Hodgkin’s Lymphoma. 2019.

